# A Pilot Study of the EMPOWER Music-based Intervention to Reduce Pulmonary Air Trapping in COPD

**DOI:** 10.64898/2026.05.26.26350616

**Authors:** Jasmine M. Taylor, Jiwoong Choi, Asma Abdolijomoor, Melissa C. Brunkan, Amy L. Wilson, Mario Castro, Nancy H. Stewart, Deanna Hanson-Abromeit, Rebecca J. Lepping

## Abstract

**Rationale:** Air trapping in functional areas of the lung is common in chronic obstructive pulmonary disease (COPD). We developed a novel music-based intervention, Engagement of Music for Pulmonary Obstruction With Expiratory Restoration (EMPOWER) aimed at reducing air trapping and functional small airways disease (fSAD) in patients with COPD.

**Objectives:** We conducted a pilot study to assess if air trapping and fSAD in COPD patients are reduced by our targeted EMPOWER music-based singing intervention.

**Methods:** Participants completed four weeks of singing and vocalizing with a board-certified music therapist. Pre- and post-intervention assessments of standard pulmonary function tests (PFTs), and quantitative computed tomography (qCT) lung imaging documented changes in air trapping. Pre- and post-intervention change in psychological and patient-reported outcomes of hope, emotional wellbeing, agency and COPD symptom burden were also obtained.

**Main Results:** All five adult participants with COPD who enrolled completed the study and reported strong interest in continuing with a similar program. Additionally, we observed trends toward improvement in qCT-measured fSAD, six-minute walk distance, and patient-reported symptoms on the COPD Assessment Test.

**Conclusion:** Results of this preliminary study showed improvements in both patient-reported and imaging respiratory outcomes, suggesting that targeted singing components in music-based interventions such as the EMPOWER intervention may support physiological lung function changes in COPD patients.

## Introduction

Chronic obstructive pulmonary disease (COPD) is classified as the fourth leading cause of mortality worldwide, with an estimate of 80 million globally having moderate to severe COPD [1] and a projected 600 million affected COPD patients by 2050 [2]. COPD causes chronic airflow obstruction and hyperinflation of the lungs, symptoms which are associated with dyspnea and decreased quality of life [3]. Air trapping in functional small airways disease (fSAD) is a common finding in COPD that is associated with dyspnea and is burdensome for patients. Previous music-based intervention studies support the use of group-based singing (i.e., choirs) to improve social and emotional outcomes for people with COPD, but there is a gap in literature that addresses the potential physiological advantages of tailored vocalization exercises to improve lung function and reduce respiratory symptoms in patients with COPD.

Because group singing activities have shown positive social and emotional improvements in patients with COPD [4-6], we further hypothesized that targeted vocal sessions that support complete exhalation may alleviate air trapping symptoms. We developed the *Engagement of Music for Pulmonary Obstruction with Expiratory Restoration* (EMPOWER) novel music-based intervention with this goal. We conducted a feasibility pilot study to measure initial patient interest and tolerability of the music-based intervention, and to look for initial evidence of physiological target engagement of reduced air trapping and functional small airways disease (fSAD) in COPD patients with the EMPOWER intervention.

The primary aim of this pilot study was to determine whether the EMPOWER music-based intervention showed promise to change respiratory outcomes. Our secondary aim was to determine the feasibility of implementation of the intervention via telehealth, use of traditional respiratory measures and the use of lung CT scans to measure change in respiratory outcomes. We hypothesized the EMPOWER music-based intervention [7] would improve respiratory exhalation and decreased air trapping mediated through progressively more complex features of pitch range, melodic contour, phrase length, tempo and timbre delivered with a tailored singing strategy[8-10]. We incorporated improvements in psychological health by promoting hope, goal adaptation, and maintaining agency [10-12]. Our goal was to directly target air trapping in regions affected by small airways disease to increase patients’ capacity to alleviate trapped air [13].

## Methods

### Study Oversight

This feasibility pilot study was conducted at a large Midwestern United States academic medical center. The study was conducted in accordance with The Common Rule (45 CFR 46) and was approved by the Institutional Review Board (IRB). The IRB protocol number is STUDY00160422. ClinicalTrials.gov Identifier: NCT07563283. Study overviews were provided to participants in person. Informed e-consent was obtained from all enrolled participants in person.

### Participants

Adults aged 18 years or older with a diagnosis of COPD were recruited from outpatient clinics at the medical center to participate in this study. Recruitment was led by the study team pulmonologist and clinic director (Author 7 – blinded for review). Eligibility for inclusion in this study required a diagnosis of COPD with airflow obstruction (FEV1/FVC < 0.7 post bronchodilator) as per GOLD guidelines [14]. Participants must have been able to hear within normal range without or with correction. An interpreter was available for eligible participants who were non-English speakers; those unable to provide consent due to cognitive impairment could participate with a study partner who could provide surrogate consent. Patients with alpha-1 antitrypsin, an inherited disorder in younger individuals causing bullous emphysema similar to COPD, and pregnant patients were excluded. Participants were compensated for their participation in study activities.

### EMPOWER Music-Based Intervention

#### Virtual Music Therapy Sessions

The EMPOWER Music-Based Intervention (MBI) was developed from a theory of intervention framework [15] and guided by intervention mapping of health need determinants, specified targeted outcomes and the Therapeutic Function of Music Plan, which defines the theory of music within a health intervention. The EMPOWER MBI has seven components that address individual respiratory change and constructs of hope with increasing complexity of primary music elements, specifically tempo, pitch, melodic contour, phrase length and timbre to facilitate targeted respiratory outcomes. The primary strategy for delivering the intervention was tailored singing that assessed participants’ current respiratory capacity and scaffolded the primary music elements to change across time. Intervention reporting details for EMPOWER are outlined using the Reporting Guidelines for Music-based Interventions checklist (Supplemental Table 1) [16-18]. EMPOWER was delivered individually to participants via HIPAA compliant telehealth (i.e., Zoom for Healthcare) by one of two board-certified music therapist who were trained on the intervention. Sessions were held virtually twice a week for four weeks, with each session lasting approximately 30-40 minutes. Participants shared their EMPOWER experience perspectives in guided, weekly diaries through verbal responses with their music therapist. The weekly diary prompts contained open-ended questions related to their perceived breathing during typical activity (conversation, moving) and singing during intervention sessions, participation goal, intervention demand (continue or recommend the intervention), and feedback on the intervention. Music therapists made brief notes of participant responses and contributions to intervention components on corresponding field notes. MT-BCs delivery fidelity was self-reported at ≥ 80% using a quality assurance checklist of session components at each visit. The session recordings were saved for implementation assessment and further operationalization of the intervention manual.

**Table 1.** Participant demographics and baseline pulmonary function. Abbreviations: BMI = body mass index; CAT = COPD Assessment Test; SGRQ = St. George’s Respiratory Questionnaire; 6MWT = six-minute walk test; DLCO = diffusing capacity of the lungs for carbon monoxide; RV = residual volume; TLC = total lung capacity; RV/TLC = residual volume/total lung capacity; FEV1 = forced expiratory volume in one second; qCT = quantitative computed tomography.

| Characteristic | Mean ± SD | Range |
| --- | --- | --- |
| Age, years | 73.60 ± 9.86 | 59-86 |
| Gender, *n* (%) female | 2 (40%) |  |
| Height, cm | 176.29 ± 85.14 |  |
| BMI, kg/m<sup>2</sup> | 25.35 ± 3.38 |  |
| Race and ethnicity |  |  |
| White, *n* (%) | 4 (80%) |  |
| Black, *n* (%) | 1 (20%) |  |
| CAT | 17.80 ± 5.68 | 8-22 |
| SGRQ Total | 39.38 ± 15.87 | 22.16-54.29 |
| SGRQ Symptoms Subscale | 54.03 ± 15.47 | 35.98-73.88 |
| SGRQ Activity Subscale | 47.22 ± 29.66 | 15.22-76.15 |
| SGRQ Impairment Subscale | 29.70 ± 13.04 | 11.89-44.88 |
| 6MWT (m) | 329.40 ± 120.96 | 149-475 |
| DLCO (ml/min/mmHg) (% predicted) | 64.20 ± 32.09 | 40-113 |
| RV (L) (% predicted) | 120.80 ± 58.75 | 55-188 |
| TLC (L) (% predicted) | 92.40 ± 24.48 | 62-114 |
| RV/TLC (L) (% predicted) | 121.20 ± 35.16 | 82-169 |
| FEV<sub>1</sub>/FVC (% predicted) | 72.60 ± 17.84 | 45-89 |
| qCT Emphysema % | 7.48 ± 9.20 | 0.20-17.60 |
| qCT fSAD % | 18.76 ± 19.15 | 1.60-41.90 |
| qCT Air Trapping % | 27.66 ± 27.61 | 3.10-60.80 |

### Questionnaires

Questionnaires of patient-reported symptoms were administered to participants within one week before and after the 4-week EMPOWER music-based intervention sessions. Patient-reported COPD questionnaires included the COPD Assessment Test [1] and St. George’s Respiratory Questionnaire [19]. Participants also completed the Adult Trait Hope Scale [8, 9]; outcomes of this measure are reported elsewhere [10]. Questionnaires were recorded and managed using REDCap electronic data capture tools [20, 21].

#### COPD Assessment Test (CAT)

The CAT Assessment contains eight patient-completed questions to assess and monitor COPD symptoms. The questionnaire measures the impact on daily life to provide a standardized measure of COPD health in patients. Participants answered questions covering cough, phlegm, breathlessness, sleep, confidence, and energy. Scores ranged from 0-5 to indicate severity with higher scores indicating worse respiratory symptoms.

#### St. George’s Respiratory Questionnaire (SGRQ)

The SGRQ contains 50 self-assessed questions divided into three main sections (Symptoms, Activity, and Impact) to assess respiratory-related quality of life. Participants answered questions to gauge frequency and severity of cough and breathlessness, physical activity limitations, and effects on daily life. Scores ranged from 0-100 with higher scores indicating maximum impairment and worse health status.

#### Adult Trait Hope Scale (AHS)

The AHS is a 12-item questionnaire to measure an individual’s general level of hope. Based on Snyder’s theory that hope involves agency and pathways. Participants responded to questions designed to gauge life outlook and their ability to achieve goals. Possible scores ranged from 8-64. Higher scores indicated greater hope.

### Pulmonary Function Test Measures

The Pulmonary function tests (PFT) used standard clinical evaluation and included pre- and post-bronchodilator measures. Metrics obtained included Forced Expiratory Volume in 1 second (FEV_1_), Forced Vital Capacity (FVC), Total Lung Capacity (TLC), Residual Volume (RV), Diffusing Capacity for Carbon Monoxide (DLCO), and Residual Volume/Total Lung Capacity (RV/TLC) ratio [22]. Lung function data are presented as percent (%) predicted based on Global GLI (i.e., non-race) predicted equations [23]. Clinical measures included the Six-Minute Walk Test (6MWT) [24], and Borg Dyspnea Index (BDI) [25] using ATS standards [26, 27].

#### Borg Dyspnea Index

The Borg Dyspnea Index (BDI) is a self-reported scale, using a 0–10-point visual-analog scale to assess patients’ perceived shortness of breath during physical activity, with 0 being no breathlessness and 10 requiring maximal effort. The BDI was collected before and after the 6MWT.

### CT Imaging Measures

CT imaging of the lungs was captured at the pre- and post-intervention visit. CT images were obtained after bronchodilator use, utilizing standardized protocols [11]. Images were collected on a Siemens CT scanner with 212 sagittal slices (2 mm slice thickness, in-plane resolution = 0.6 × 0.6 mm) for inspiratory and expiratory states. We conducted a quantitative CT (qCT) analysis to quantify fSAD at baseline and follow-up post intervention. Airway, lung, and lobe segmentation and quantitative analyses were done with Apollo 2.0/Vision 2.2 (VIDA Diagnostics, Inc., Coralville, Iowa) and *in-house* software.

### Data Processing

#### qCT Analysis

Individual inspiratory and expiratory image series were imported in VIDA Vision 2.2 software (VIDA Diagnostics, Coralville, IA, USA) and processed for segmentation of airway, lungs, lobes, and pulmonary vessels, and measurements of sizes and CT densities. Then, an in-house qCT pipeline was used for multiscale (whole and regional) lung structure-function analysis, including air trapping percentage (AirT%) and functional small airways disease percentage (fSAD%).

#### Statistical Analysis

Separate paired sample t-tests were used to compare individual participant change in each clinical, questionnaire, and imaging measure pre-to post-intervention. Statistical significance threshold for each test was set at two-sided *p* < .05. For this exploratory pilot study, no correction was made for multiple comparisons. Mean differences and effect sizes (Cohen’s *d*) are reported. Analyses were conducted in IBM SPSS for Macintosh, version 29.0.0 (IBM Corp., Armonk, N.Y., USA).

## Results

### Participants

Seven adults with COPD were screened for eligibility. After screening, two eligible individuals declined to participate. One declined due to transportation limitations. The second eligible person declined due to a conflicting schedule. Five participants enrolled and completed all pre-intervention assessments, the 8 session EMPOWER music-based intervention and post-intervention assessments. Demographics and baseline lung function characteristics are reported in Table 1.

### Pulmonary Function Test Measures

#### Six-minute walk test

Although not statistically significant (*p* = .51, *d* = -0.32), participants walked an average of 28.10 m further following the EMPOWER MBI (Table 2).

**Table 2.** Respiratory and clinical outcomes. Abbreviations: df = degrees of freedom; 6MWT = six-minute walk test; m = meters; BDI = Borg Dyspnea Index; DLCO = diffusing capacity of the lungs for carbon monoxide; RV/TLC = residual volume/total lung capacity; FEV1 = forced expiratory volume in one second; BD = bronchodilator. * indicates *p* < .05. ** indicates *p* < .01

| Measure | Pre<br>(Mean) | Post (Mean) | Mean<br>Difference<br>(SD<sub>DIFF</sub>) | t | df | p<br>(two<br>sided) | Effect<br>size<br>(Cohen's<br>d) |
| --- | --- | --- | --- | --- | --- | --- | --- |
| 6MWT (m) | 329.40 | 357.50 | -28.10<br>(87.56) | -0.72 | 4 | .51 | -0.32 |
| BDI (Before walk) | 2.43 | 2.00 | 0.43 (0.51) | 1.46 | 2 | .28 | 0.84 |
| BDI (After walk) | 2.33 | 2.95 | -0.63<br>(0.95) | -1.32 | 3 | .28 | -0.66 |
| DLCO<br>(ml/min/mmHg)<br>(% predicted) | 64.20 | 59.40 | 4.80 (2.39) | 4.50 | 4 | .01** | 2.01 |
| RV/TLC (L) (%<br>predicted) | 121.20 | 121.60 | -0.40<br>(12.99) | -0.07 | 4 | .95 | -0.03 |
| FEV<sub>1</sub> (Pre BD) (L)<br>(% predicted) | 55.20 | 53.00 | 2.20 (2.56) | 1.90 | 4 | .13 | 0.85 |
| FEV<sub>1</sub> (Post BD) (L)<br>(% predicted) | 59.20 | 55.80 | 3.40 (5.37) | 1.42 | 4 | .23 | 0.63 |

#### Borg Dyspnea Index (BDI)

BDI did not change (*p* = .28, *d* = 0.84; Table 2).

#### Pulmonary Function Test (PFT)

All PFT outcome variables (FEV_1_, FVC, TLC, RV, RV/TLC ratio, DLCO, 6MWT, and BDI) were not significantly changed following the EMPOWER MBI, with the exception of DLCO, which had a 4.8% worsening (64.20% to 59.40%). This change was statistically significant (*p* =.01) in this small sample; however, this is not a clinically meaningful difference (Table 2).

### Patient-Reported Outcome Measures

#### COPD Assessment Test (CAT)

The total score of the CAT decreased by 2 points, which is a clinically meaningful difference. This change did not reach statistical significance (*p* = .08, *d* = 1.07) (Table 3).

**Table 3.** Patient-reported outcomes. Abbreviations: df = degrees of freedom; CAT = COPD Assessment Test; SGRQ = St. George’s Respiratory Questionnaire. * indicates *p* < .05. ** indicates *p* < .01.

| Measure | Pre<br>(Mean) | Post<br>(Mean) | Mean<br>Difference<br>(SD<sub>DIFF</sub>) | t | df | p<br>(two<br>sided) | Effect<br>size<br>(Cohen's<br>d) |
| --- | --- | --- | --- | --- | --- | --- | --- |
| **CAT Total** | **17.80** | **15.80** | **2.00 (1.87)** | **2.39** | **4** | **.08** | **1.07** |
| *CAT Sleep* | *2.20* | *1.60* | *0.60 (0.89)* | *1.50* | *4* | *.21* | *0.67* |
| *CAT Energy* | *2.80* | *3.00* | *-0.20 (0.45)* | *-1.00* | *4* | *.37* | *-0.45* |
| **SGRQ Total** | **39.38** | **38.59** | **0.79 (11.22)** | **0.16** | **4** | **.88** | **0.07** |
| *SGRQ*<br>*Symptoms* | *54.03* | *52.50* | *1.53 (18.01)* | *0.19* | *4* | *.86* | *0.09* |
| *SGRQ*<br>*Activities* | *47.22* | *56.36* | *-9.15 (14.39)* | *-1.42* | *4* | *.23* | *-0.64* |
| *SGRQ*<br>*Impairment* | *29.70* | *23.25* | *6.45 (10.46)* | *1.38* | *4* | *.24* | *0.62* |
| **Hope Total** | **46.60** | **49.40** | **-2.80 (6.80)** | **-0.92** | **4** | **.41** | **-0.42** |
| *Hope Agency* | *22.00* | *24.00* | *-2.00 (4.95)* | *-0.90* | *4* | *.42* | *-0.40* |
| *Hope Pathway* | *24.60* | *25.40* | *-0.80 (2.68)* | *-0.67* | *4* | *.54* | *-0.30* |

#### St. George’s Respiratory Questionnaire (SGRQ)

There was considerable variability between participants on SGRQ change, with no significant change with the program (*p* = .88, *d* = 0.07; Table 3).

#### Adult State Hope Scale

Hope scores increased by 2.8 points, which was not statistically significant (*p* = .41, *d* = - 0.42) (Table 3).

### CT Imaging Measures

As the primary analysis of changes in air trapping and fSAD between pre- and post-EMPOWER MBI sessions, AirT% and fSAD% at the whole lung were compared between the two visits. Both AirT% and fSAD% had consistent improvements across the group; however, these improvements failed to reach statistical significance (*p* = .09, *d* = 0.98-1.00) (Table 4). This was due to variability in baseline air trapping between participants. Figure 1 provides a visualization of the patient variability in air trapping change. Three of the five participants had noticeably high AirT% (61%, 54%, and 17% of the whole lung). After the singing sessions, AirT% was decreased in all three by 1.3%, 15%, and 3.2% in the whole lung. The other two had baseline AirT% as low as 4% and 3%, which remained low with changes less than 1% after participating in the EMPOWER MBI sessions. One participant with AirT% of 61% in the whole lung had 54% in the right upper lobe (RUL), where AirT% decrease by 4.5% after singing. Another participant, who had AirT% of 54% also showed the largest reduction of AirT% in the RUL (baseline 57%, reduction by 19%). Both were the patients with noticeable emphysema score by inspiratory low attenuation area percent below -950 HU (LAA_IN_%) of 18% (both), which were also reduced (1.7% and 12.4%, respectively). The other three had negligible baseline emphysema score less than 2% of the whole lung. One participant had noticeable AirT% of 18% only in the right middle lobe (RML). fSAD% showed similar trends.

**Table 4.** qCT lung function outcomes. Abbreviations: df = degrees of freedom; fSAD = functional small airways disease; AirT = air trapping. * indicates *p* < .05. ** indicates *p* < .01.

| Measure | Pre<br>(Mean) | Post<br>(Mean) | Mean<br>Difference<br>(SD<sub>DIFF</sub>) | T | df | p<br>(two<br>sided) | Effect<br>size<br>(Cohen's<br>d) |
| --- | --- | --- | --- | --- | --- | --- | --- |
| Emphysema% | 7.48 | -2.86 | 10.34 (13.58) | 1.70 | 4 | .16 | 0.76 |
| fSAD% | 18.76 | -1.82 | 20.58 (20.61) | 2.23 | 4 | .09 | 1.00 |
| AirT% | 27.66 | -3.68 | 31.34 (31.84) | 2.20 | 4 | .09 | 0.98 |

**Figure 1.**
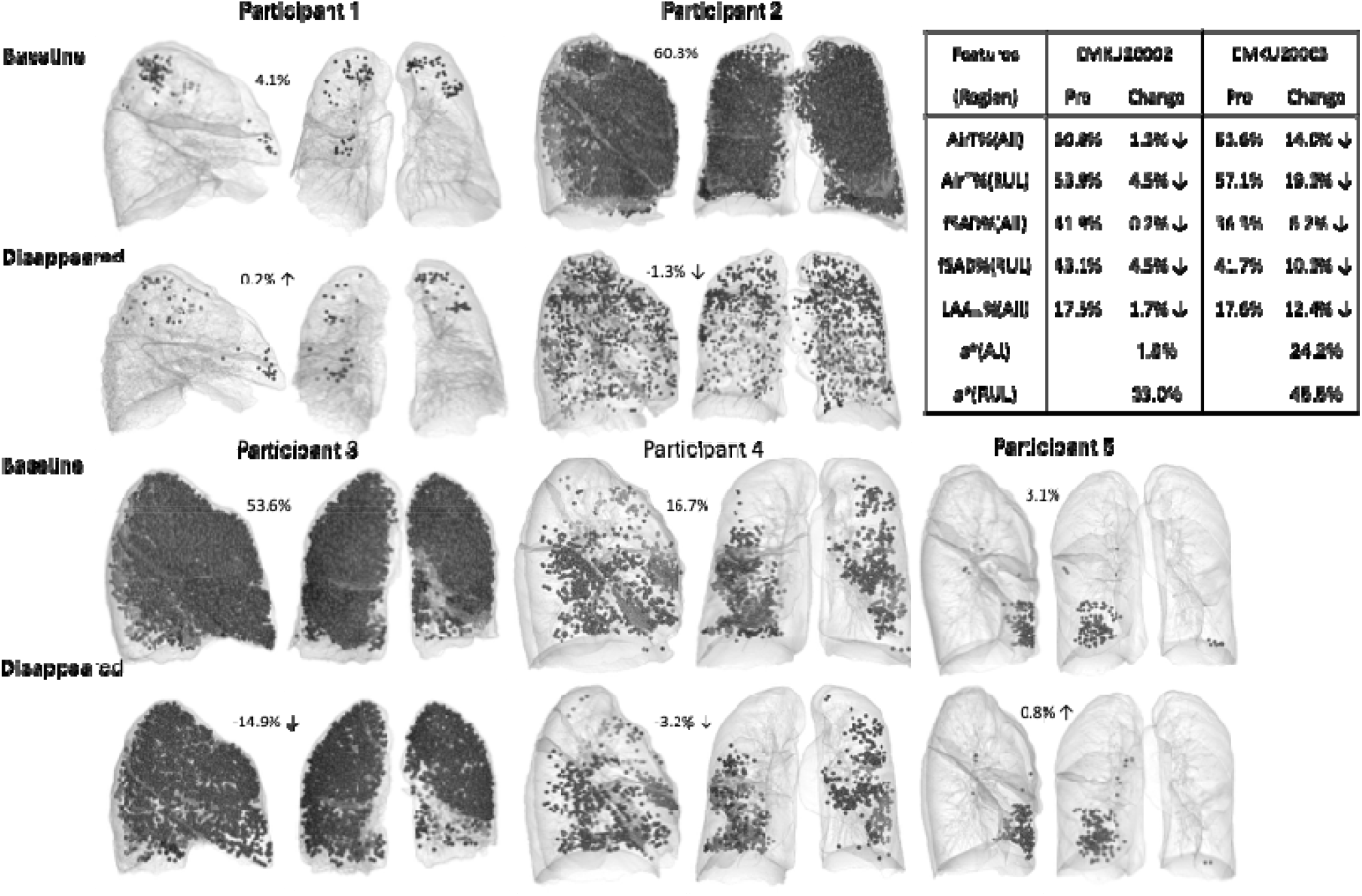
qCT lung maps of air trapping of all five participants and selected features of two participants.

### Safety and tolerability

COPD exacerbations were not observed in any of the five participants. One participant experienced an unrelated hospitalization after the first EMPOWER MBI session and completed the remaining seven sessions upon hospital discharge. All five participants completed the twice a week, eight session EMPOWER MBI and post-intervention assessments.

## Discussion

In this study, we developed the EMPOWER music-based intervention to target respiratory function for people with COPD. We conducted a pilot feasibility and preliminary target-engagement study of the impact of our EMPOWER intervention on clinical and imaging pulmonary function outcomes. Our study was well-tolerated by patients, as indicated by 100% completion by all enrolled participants, no pulmonary exacerbations, and all participants reporting that they enjoyed the program and would like to continue with a similar intervention.

We also observed small, but promising changes in clinical outcomes and meaningful improvements on our primary imaging outcome of fSAD%. After the eight sessions of the EMPOWER MBI, all three participants with noticeable air trapping at baseline experienced reduction of AirT% and fSAD%. LAA_IN_% that indicates degrees of emphysema or inspiratory hyperinflation was also reduced in the two participants with noticeable LAA_IN_%. The use of sensitive qCT imaging markers of lung function alongside clinically interpretable outcomes is a strength of our study design.

Prior studies of singing for COPD have primarily involved group-based singing, which has shown consistent improvements for mood, wellbeing, and quality of life. This is an important aspect of COPD management, as the burden of respiratory symptoms often leads to anxiety, isolation, loneliness and depression. These group singing interventions focus on the social components of music-making; however, there is an assumption that the act of singing and exercising the respiratory system would improve lung function outcomes. The literature is mixed on respiratory outcomes, potentially due to variability in the effective delivery of targeted musical components that have the potential to shift specific symptomatic targets. Our EMPOWER MBI was designed specifically to target full respiratory expiration and is a key strength of our music-based intervention. Another key strength is the use of lung CT scans as a measure sensitive to changes.

Prior to the implementation of this study a targeted intervention did not exist and had to be developed. Our EMPOWER intervention was operationalized and designed by board-certified Music Therapists, skilled music-based interventionists, and vocal production experts, utilizing the Therapeutic Function of Music Plan framework [17, 18, 28]. The EMPOWER manual informed the intervention delivery fidelity criteria. Interventionists who provided the EMPOWER sessions were masters level and experienced board-certified Music Therapists who were trained on the intervention. During training their input further refined the intervention for implementation.

The intervention manual, corresponding session quick reference outlines, participant guides, and delivery checklists were provided for each Music Therapist. Materials were individualized for each participant to contain their chosen anchor song from a pre-selected list of common songs known to be preferred by people within the age range of the majority of people diagnosed with COPD.

### Limitations

This feasibility pilot study has several limitations, and the results should be interpreted cautiously. Notably, the sample size is very small, with only five participants in this preliminary feasibility and target-engagement study. These five participants also had variability in some baseline measurements. Severity of air trapping, emphysema and hyperinflation varied between participants. Comprehensive regional lung structure-function analysis with more cases may help derive associated mechanisms. More data are required for appropriate interpretation and statistical analysis of the findings. For a better understanding of the mechanism of air trapping reduction, more comprehensive qCT and computational fluid dynamics (CFD) analysis are needed, which were not conducted here due to the limited sample size and the risk of overinterpretation. Additionally, our qCT data were conducted under typical clinical assessment at rest and not during singing or vocalization. Future studies should conduct CFD analysis under individualized breathing conditions during singing sessions to provide the most accurate analysis. Studies could also include data from handheld spirometers or similar flow measurement devices to collect valuable breathing waveform data. To better interpret the impact of air trapping reduction for daily symptom management, measurement of daily normal breathing (e.g., tidal volume and respiratory rate) could provide valuable information for in-depth CFD analysis.

It should also be noted that the lack of control group, participant self-selection into the study, lack of randomization, and lack of blinding also limit the interpretation of the findings. Future research must include randomized controlled trials with larger and more diverse samples to clearly estimate the impact of the EMPOWER music-based singing intervention of lung function in patients with COPD.

### Conclusion

In conclusion, we have developed a targeted music-based intervention aimed at reducing air trapping in functional small airways for people with COPD. Our EMPOWER intervention yielded small, but promising improvements in clinical outcomes, patient-reported symptoms, and sensitive qCT imaging markers of target engagement. The EMPOWER intervention shows promise as a tailored music-based intervention for people with COPD and contributes to the current literature on music-based interventions that have well-documented impacts on psychological wellbeing and quality of life.

## Data Availability

All data produced in the present study are available upon reasonable request to the authors.

## Acknowledgements

The authors thank Zaid M. Mansour, Ph.D. for his contributions to the development of this work, Jennifer Welch, MMT, MT-BC, SEP, CHt for her contributions as a music interventionist for the virtual sessions, David Lee for his contributions to data processing, and Benjamin Hess for his contributions to the project. We humbly thank the research participants for taking part in this preliminary investigation and for their valuable feedback on the development of the intervention.

## Notes

**Funding:** This work was supported by the KUMC Division of Pulmonology, Department of Internal Medicine, Department of Neurology, Hoglund Biomedical Imaging Center, and the University of Kansas School of Music. Dr. Lepping was supported by the National Center for Advancing Translational Sciences of the National Institutes of Health under the Award Number UL1TR002366. The contents are solely the responsibility of the authors and do not necessarily represent the official views of the NIH or NCATS.

### Competing Interest Statement

The authors have declared no competing interest.

### Clinical Trial

NCT07563283

### Funding Statement

This work was supported by the KUMC Division of Pulmonology, Department of Internal Medicine, Department of Neurology, Hoglund Biomedical Imaging Center, and the University of Kansas School of Music. Dr. Lepping was supported by the National Center for Advancing Translational Sciences of the National Institutes of Health under the Award Number UL1TR002366. The contents are solely the responsibility of the authors and do not necessarily represent the official views of the NIH or NCATS.

### Author Declarations

The IRB of the University of Kansas Medical Center gave ethical approval for this work.

