## Supplemental Table 1 for "A Pilot Study of the EMPOWER Music-based Intervention to Reduce Pulmonary Air Trapping in COPD"

Reporting Guidelines for Music-based Interventions checklist[^a^](https://pmc-ncbi-nlm-nih-gov.kumc.idm.oclc.org/articles/PMC12171218/table/tab1/#tfn1).

| **Item number** | | **Item** | **Location**[**^b^**](https://pmc-ncbi-nlm-nih-gov.kumc.idm.oclc.org/articles/PMC12171218/table/tab1/#tfn2) **(page or appendix number)** |
| --- | --- | --- | --- |
| 1 | | **Brief Name**[^c^](https://pmc-ncbi-nlm-nih-gov.kumc.idm.oclc.org/articles/PMC12171218/table/tab1/#tfn3) Provide the name or phrase that describes the intervention. | Abstract |
| 2 | | **Intervention Theory and/or Scientific Rationale** Provide a rationale for the music and/or music experience(s). Specify how essential features of the music and music experience(s) are expected to influence targeted outcomes. | Page 3 |
| 3 |  | **Intervention Content** For Items 3a–3e, describe the music intervention with enough detail to support replication. When applicable, describe procedures for tailoring the intervention. | Page 8 |
|  | 3a | **Music Selection** Describe the process for how music was selected including who was involved in music selection. | Page 8 |
|  | 3b | **Music** Specify key details about the music that may be relevant to specified outcomes of interest. Characteristics may include compositional features of the music (such as tempo, harmony, rhythm, pitch, tonality, form, instrumentation),[^d^](https://pmc-ncbi-nlm-nih-gov.kumc.idm.oclc.org/articles/PMC12171218/table/tab1/#tfn4) sound intensity or volume, lyrics, and/or how the music relates to the participants’ cultural identity and heritage. When using published music, provide reference for a sound recording or sheet music. | Page 8 |
|  | 3c | **Music Delivery Method** Provide details about how music was provided to or created with participants (such as live, recorded, computer generated).[^d^](https://pmc-ncbi-nlm-nih-gov.kumc.idm.oclc.org/articles/PMC12171218/table/tab1/#tfn4) Include any details necessary for replication. This might include size of performing group, use of playback equipment, or person controlling volume. | Page 8 |
|  | 3d | **Materials** List all materials necessary for the music experience. Include music and non-music equipment and materials. | Page 8 |
|  | 3e | **Intervention Strategies** Describe the music intervention strategy or strategies being studied (such as music listening, improvisation, song writing, rhythmic auditory stimulation).[^d^](https://pmc-ncbi-nlm-nih-gov.kumc.idm.oclc.org/articles/PMC12171218/table/tab1/#tfn4) | Page 8 |
| 4 | | **Interventionist** Specify interventionist qualifications, credentials, training, and/or experience. Indicate how many interventionists delivered the music experience. | Page 8 |
| 5 | | **Individual or Group Intervention** Specify whether interventions were delivered to individuals or groups of individuals. For group interventions, specify the size of the group. | Page 8 |
| 6 | | **Setting** Describe where the intervention was delivered. Include location, privacy level, ambient sound, and/or any other factors that may have affected participants’ experiences. | Page 8 |
| 7 | | **Intervention Delivery Schedule** Report number of sessions, session length (for example, 60 min), frequency (for example, 3×/week), time interval between sessions (for example, single day, three consecutive days), and duration (for example, over 4 weeks).[^d^](https://pmc-ncbi-nlm-nih-gov.kumc.idm.oclc.org/articles/PMC12171218/table/tab1/#tfn4) Include practice, experiences, or tasks that are assigned to participants between intervention sessions. | Page 8 |
| 8 | | **Treatment Fidelity** Describe strategies and/or measures used to ensure that the music intervention was delivered and received as intended. | Page 8 |
